# School immunization coverage during the COVID-19 pandemic: A retrospective cohort study

**DOI:** 10.1101/2022.05.04.22274665

**Authors:** Hannah Sell, Yuba Raj Paudel, Donald Voaklander, Shannon E. MacDonald

**Affiliations:** School of Public Health, University of Alberta, Edmonton, Alberta, Canada; Faculty of Nursing, University of Alberta, Edmonton, Alberta, Canada

**Keywords:** vaccine, immunization, HPV, Meningococcal, COVID-19, coverage, pandemic, adolescent, school

## Abstract

Few studies have assessed the impact of the COVID-19 pandemic on immunization coverage for adolescents, and little is known about how coverage has changed throughout the pandemic. We aimed to: (1) assess the change in coverage for school-based vaccines in Alberta, Canada resulting from the pandemic; (2) determine whether coverage differed by geographic health zone and school type; and (3) ascertain whether coverage has returned to pre-pandemic levels. Using a retrospective cohort design, we used administrative health data to compare coverage for human papillomavirus (HPV) and meningococcal conjugate A, C, Y, W-135 (MenC-ACYW) vaccines in Alberta, Canada between pre-pandemic (2017-2018 school year) and pandemic (2019-2020 and 2020-2021 school years) cohorts (N=289,420). Coverage was also compared by health zone and authority type. The 2019-2020 cohort was followed over one year to assess catch-up. Compared to 2017-2018, immunization coverage for HPV was significantly lower in the 2019-2020 (absolute difference: 60.8%; 95% CI: 60.4-61.3%) and 2020-2021 cohorts (absolute difference: 59.9%; 95% CI: 59.4-60.3%). There was a smaller, significant decline in MenC-ACYW coverage comparing 2017-2018 to 2019-2020 (absolute difference: 6.1%; 95% CI: 5.6-6.5%) and 2020-2021 (absolute difference: 32.2%; 95% CI: 31.6-32.7%). Private schools had low coverage overall, while coverage fluctuated by zone. During follow-up of the 2019-2020 cohort, coverage for HPV and MenC-ACYW increased from 5.6% to 50.2%, and 80.7% to 83.0%, respectively. There was a substantial decrease in school-based immunization coverage during the COVID-19 pandemic, and coverage has not returned to pre-pandemic levels, suggesting further catch-up is needed.

## Introduction

The COVID-19 pandemic has greatly impacted health care systems worldwide via COVID-19-related hospitalizations, testing and tracing programs, and the introduction of mass COVID-19 immunization campaigns. As a result, many routine health services may have been overlooked, including immunization. There has been mounting concern among public health experts and health care providers regarding global declines in routine childhood immunization coverage during the pandemic.^1^

In Canada, public health restrictions began in mid-March 2020, including physical distancing measures, school and clinic closures, and re-allocation of public health staff.^4^ These restrictions may have disrupted routine immunization programs, leaving many Canadian children with missed immunization doses.^5,6^ Immunizations for school-aged children have likely been significantly disrupted as national recommendations have prioritized infant vaccines, while school-based immunization programs were suspended due to school closures.^5,7^ As such, there is concern that the reduction in coverage for adolescent vaccines will increase susceptibility to vaccine-preventable diseases.^5,8^

Few studies have assessed the impact of the COVID-19 pandemic on immunization coverage for adolescents. For instance, two U.S. studies reported significant declines in immunization rates for adolescents in the early pandemic period (March-May 2020).^2,3^ However, no studies to date have examined how coverage for adolescent vaccines has changed throughout the pandemic. It is essential to quantify the impact of the pandemic on routine immunization coverage and determine whether coverage recovered during the pandemic, as this information can assist public health in identifying populations of children with missed immunizations and devising strategies for catch-up to prevent future disease outbreaks. As such, the objective of this study was to assess the change in immunization coverage resulting from the COVID-19 pandemic for two school-based vaccines (human papillomavirus [HPV] and meningococcal A, C, Y, W-135 [MenC-ACYW]) and ascertain whether coverage has returned to pre-pandemic levels. Additionally, we sought to determine whether immunization coverage during the pandemic differed by factors such as geographic health zone and school type.

## Methods

### Study design

This was a retrospective cohort study of immunization coverage for two school-based vaccines using administrative health data.

### Setting

The study took place in Alberta, a western Canadian province with a population of approximately 4.5 million.^9^ The province is divided into five geographical health zones (North, South, and Central, which are predominantly rural zones, and Calgary and Edmonton, which are large urban zones) for the purpose of publicly funded health service delivery, including immunizations.^10^ Routinely scheduled immunizations for adolescents are usually administered by public health nurses through school-based programs.^11^ Table A1 in the appendix provides more information on the number and timing of doses for school-based immunizations in Alberta.

### Data sources

The dataset was created by linking population-based administrative databases from the Alberta Ministry of Health, including Alberta Health Care Insurance Plan (AHCIP), Provincial School Immunization Record (PSIR), and Immunization and Adverse Reaction to Immunization (Imm/ARI). Almost all Albertans (99%) are registered in the AHCIP, which assigns them a unique lifetime identifier that was used to link administrative health records for each individual, including their immunization records. During the period of this study, the Imm/ARI database included all publicly funded childhood vaccines administered in Alberta, except those described in exclusions below. The PSIR database contains individual-level records of student enrollment information in each school year, including grade, age, and sex, as well as school-level information such as geographic health zone and authority type.

#### Exclusions

Exclusions were those who died or migrated out of Alberta during each respective school year, identified as First Nations (Canada’s largest population of Indigenous Peoples, who were excluded because their data is not consistently submitted to Imm/ARI and PSIR), or were from Lloydminster (as vaccines are delivered by a neighbouring province). Additionally, schools that offer online or distance education (not due to COVID-19), post-secondary, continuing education, summer, or evening/weekend schools were also excluded. To be consistent with previous coverage analyses,^12^ age limitations were also applied for each grade: for grade 5, those <9 years of age and >11 years were excluded from the analysis; for grade 6, those <10 years and >12 years were excluded; for grade 9, those <13 years and >15 years were excluded. Figures A1 and A2 in the appendix provide further details on exclusions.

### Analysis

Immunization coverage^13^ was defined as the proportion of age- and grade-eligible children who received a complete vaccine series on or prior to July 31 of each school year for two vaccines, HPV and MenC-ACYW. Currently, a complete vaccine series for HPV consists of two doses given in grade 6, while MenC-ACYW is a single dose given in grade 9. There was no school program for HPV immunization in the 2018-2019 school year due to a change in delivery from three doses in grade 5 to two doses in grade 6,^14^ so the 2017-2018 school year was used as the pre-pandemic comparison year. As a three-dose program was in the place throughout 2017-2018, receipt of at least three doses was used to define a complete HPV series for this cohort. As some students may have received more doses than required, a complete series was defined as receiving the number of required doses as a minimum. Coverage was calculated as the number of children who had received a complete dose series of HPV (at least two doses in 2019-2020 or 2020-2021 and at least three doses in 2017-2018) or MenC-ACYW (at least one dose) vaccines divided by the number of children who had a record in PSIR of attending grade 5, 6, or 9 between September 1 and July 31. Absolute differences in coverage between the pre-pandemic (2017-2018 school year) and pandemic (2019-2020 and 2020-2021 school years) cohorts for each vaccine and associated 95% confidence intervals (CIs) were calculated and compared using Pearson’s chi-square tests. Coverage was also stratified by geographic health zone (i.e., Calgary, Edmonton, Central, North, South) based on school location, and school authority type (i.e., public, publicly-funded Catholic, private, charter, francophone). Crude RRs and 95% CIs were calculated for the analysis stratified by the geographic health zone (reference: Calgary) and authority type (reference: public); the zone and authority type with the largest populations were chosen as reference categories. A small number of participants (<1%) had missing data for the zone variable but were still included in the cohorts as no other data were missing.

To assess catch-up over time for both HPV and MenC-ACYW, children who were due to be vaccinated in the 2019-2020 school year were followed until September 1, 2021. Absolute differences in coverage and 95% CIs were calculated for each vaccine after the catch-up period (September 1, 2021) in comparison to coverage for the pre-pandemic (2017-2018) cohort at July 31, 2018 (which represented coverage at the end of a ‘typical’ school year). Differences were compared using Pearson’s chi-square tests. Coverage for HPV and MenC-ACYW for the 2019-2020 cohort over the follow-up period was also stratified and presented by geographic health zone and authority type. This analysis excluded any individual from the 2019-2020 cohort who died or left the province before September 1, 2021. Individuals from the 2020-2021 cohort were not followed due to the short period of time between assessment of coverage for this cohort and the end of the follow-up period (July 31, 2021 to September 1, 2021). SAS version 9.4 (SAS Institute Inc., Cary, NC, USA) was used for all analyses. This study received approval from the Health Research Ethics Board at the University of Alberta (ethics ID: Pro00102401).

## Results

Table 1 describes the number of students and the sociodemographic characteristics of each cohort.

**Table 1.** Sociodemographic characteristics of the cohorts.

|  | **HPV cohorts (grades 5 & 6)<sup>a</sup>** |  |  | **MenC-ACYW cohorts (grade 9)** |  |  |
| --- | --- | --- | --- | --- | --- | --- |
|  | **Pre-pandemic** | **Pandemic** |  | **Pre-pandemic** | **Pandemic** |  |
| **<u>Characteristic</u>** | ***2017-2018*** | ***2019-2020*** | ***2020-2021*** | ***2017-2018*** | ***2019-2020*** | ***2020-2021*** |
| ***N*** | 50,127 | 51,760 | 52,770 | 42,442 | 45,376 | 46,945 |
| **Age, mean (SD)** | 9.70<br>(0.50) | 10.70<br>(0.49) | 10.70<br>(0.49) | 13.69<br>(0.46) | 13.68<br>(0.46) | 13.67<br>(0.47) |
| **Gender, *n* (%)** |  |  |  |  |  |  |
| Female | 24,463<br>(48.80) | 25,215<br>(48.71) | 25,624<br>(48.55) | 20,858<br>(49.14) | 22,196<br>(48.91) | 22,880<br>(48.74) |
| Male | 25,662<br>(51.19) | 26,536<br>(51.27) | 27,122<br>(51.40) | 21,581<br>(50.85) | 23,163<br>(51.05) | 23,996<br>(51.11) |
| Other | 2<br>(0.01) | 9<br>(0.02) | 24<br>(0.05) | 3<br>(0.01) | 17<br>(0.04) | 69<br>(0.15) |
| **Geographic health zone, *n* (%)** |  |  |  |  |  |  |
| Calgary | 19,734<br>(39.37) | 20,629<br>(39.86) | 21,270<br>(40.31) | 16,899<br>(39.82) | 17,920<br>(39.49) | 19,176<br>(40.85) |
| Edmonton | 16,020<br>(31.96) | 16,665<br>(32.20) | 17,136<br>(32.47) | 13,471<br>(31.74) | 14,697<br>(32.39) | 15,312<br>(32.62) |
| Central | 5,204<br>(10.38) | 5,172<br>(9.99) | 5,220<br>(9.89) | 4,529<br>(10.67) | 4,642<br>(10.23) | 4,677<br>(9.96) |
| North | 5,086<br>(10.15) | 5,249<br>(10.14) | 5,233<br>(9.92) | 4,407<br>(10.38) | 4,685<br>(10.32) | 4,624<br>(9.85) |
| South | 3,678<br>(7.34) | 3,641<br>(7.03) | 3,772<br>(7.15) | 2,809<br>(6.62) | 3,068<br>(6.76) | 3,007<br>(6.41) |
| Missing | 405<br>(0.81) | 404<br>(0.78) | 139<br>(0.26) | 327<br>(0.77) | 364<br>(0.80) | 149<br>(0.32) |
| **School authority type, *n* (%)** |  |  |  |  |  |  |
| Public | 34,175<br>(68.18) | 35,176<br>(67.96) | 35,388<br>(67.06) | 28,873<br>(68.03) | 31,074<br>(68.48) | 31,874<br>(67.90) |
| Catholic | 12,050<br>(24.04) | 12,632<br>(24.40) | 12,936<br>(24.51) | 10,814<br>(25.48) | 11,400<br>(25.12) | 11,908<br>(25.37) |
| Private | 2,253<br>(4.49) | 2,309<br>(4.46) | 2,750<br>(5.21) | 1,611<br>(3.80) | 1,727<br>(3.81) | 1,863<br>(3.97) |
| Charter | 982<br>(1.96) | 978<br>(1.89) | 988<br>(1.87) | 828<br>(1.95) | 812<br>(1.79) | 842<br>(1.79) |
| Francophone | 650<br>(1.30) | 665<br>(1.28) | 708<br>(1.34) | 315<br>(0.74) | 363<br>(0.80) | 458<br>(0.98) |

Notes. HPV = human papillomavirus; MenC-ACYW = meningococcal A, C, Y, W-135; SD = standard deviation.
<sup>a</sup>Starting in the 2018-2019 school year, there was a change in delivery for HPV vaccine from three doses given in grade 5 to two doses in grade 6, so there was no school program for HPV in the 2018-2019 school year. As such, the 2017-2018 school year was used as the pre-pandemic comparison year.

### HPV coverage

Full coverage (i.e., two or more doses in 2019-2020 and 2020-2021, three or more doses in 2017-2018) for the HPV vaccine on July 31 of each respective school year was 66.4% in the 2017-2018 pre-pandemic cohort, 5.6% in the 2019-2020 pandemic cohort, and 6.6% in the 2020-2021 pandemic cohort (Figure 1). In comparison to 2017-2018, coverage for HPV was significantly lower in 2019-2020 (5.6% vs. 66.4%; absolute difference: 60.8%; 95% CI: 60.4-61.3%; p < 0.001) and 2020-2021 (6.6% vs. 66.4%; absolute difference: 59.9%; 95% CI: 59.4-60.3%; p < 0.001).

**Figure 1.**
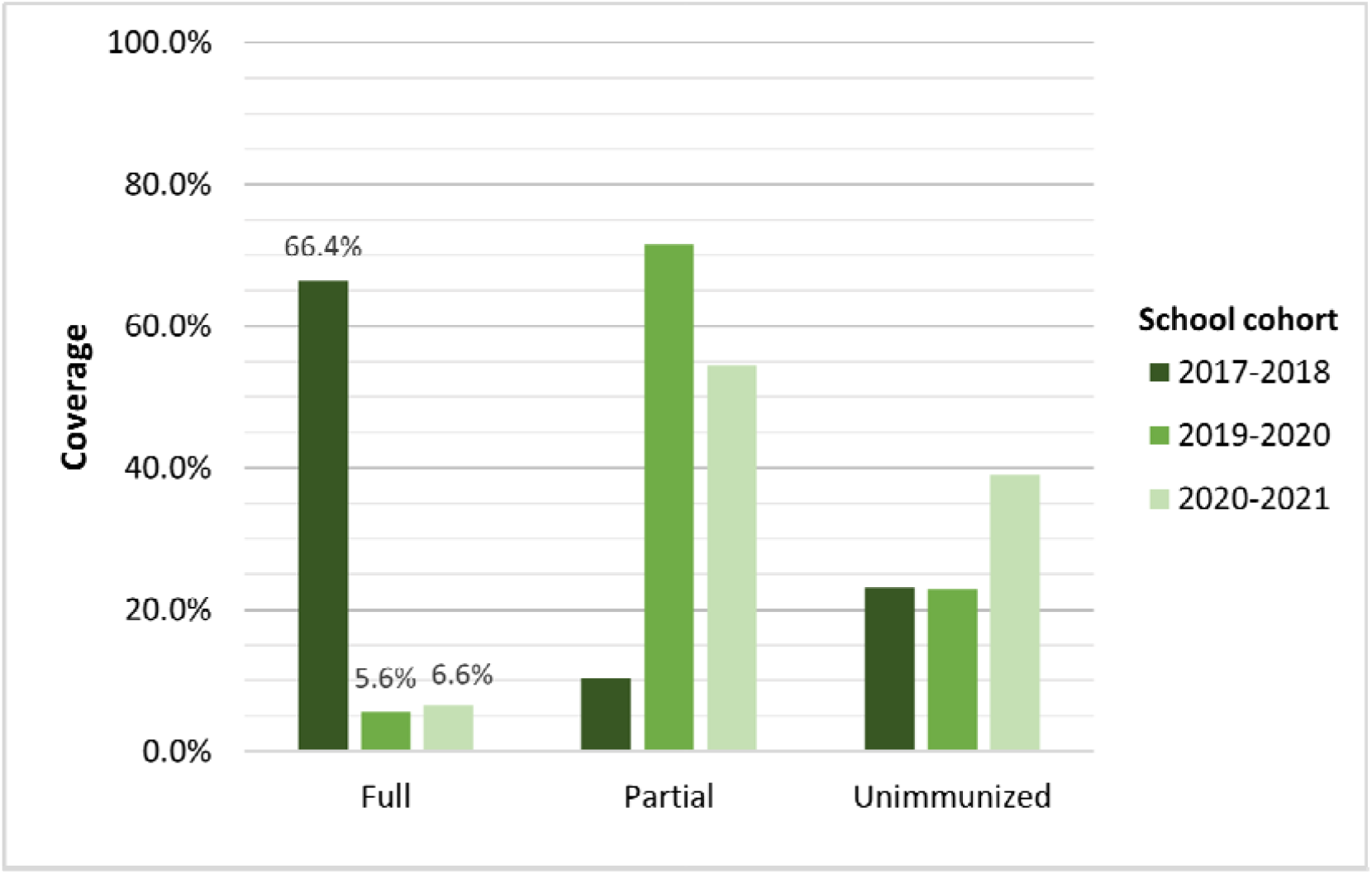
Coverage (full^a^, partial^b^, or unimmunized) for HPV vaccine in the pre-pandemic (2017-2018) and pandemic (2019-2020, 2020-2021) school cohorts at July 31 of each respective school year. *Notes*. HPV = human papillomavirus. The HPV school immunization program in Alberta changed from a three-dose schedule given in grade 5 to a two-dose schedule given in grade 6 in the 2018-2019 school year. As such, no immunization data were available for the 2018-2019 school year, so 2017-2018 was used as the pre-pandemic comparison year. ^a^Full coverage defined as the percentage of children who received all scheduled doses of HPV vaccine by July 31 of each respective school year (i.e., at least two doses for the 2019-2020 and 2020-2021 cohorts, at least three doses for the 2017-2018 cohort). ^b^Partial coverage defined as the percentage of children who received some but not all scheduled doses of HPV by July 31 of each respective school year (i.e., one dose for the 2019-2020 and 2020-2021 cohorts, one or two doses for the 2017-2018 cohort).

Table 2 shows HPV immunization coverage at July 31 of each pandemic school year by geographic health zone and school type. Across health zones, coverage ranged from 1.1% (Edmonton) to 24.7% (Central) in the 2019-2020 cohort and 0.3% (South) and 11.9% (Edmonton) in the 2020-2021 cohort. Public schools had the highest HPV coverage in both pandemic cohorts (6.6% and 7.4%), while charter and private schools had the lowest coverage in 2019-2020 and 2020-2021, respectively.

**Table 2.** Immunization coverage for HPV and MenC-ACYW vaccines in the 2019-2020 and 2020-2021 cohorts by geographic health zone and school authority type.

| <u>School year</u> | <u>HPV (grade 6)</u> |  |  |  | <u>MenC-ACYW (grade 9)</u> |  |  |  |
| --- | --- | --- | --- | --- | --- | --- | --- | --- |
|  | <u>2019-2020</u> |  | <u>2020-2021</u> |  | <u>2019-2020</u> |  | <u>2020-2021</u> |  |
|  | Coverage (%) | RR (95% CI) | Coverage (%) | RR (95% CI) | Coverage (%) | RR (95% CI) | Coverage (%) | RR (95% CI) |
| **Geographic health zone** |  |  |  |  |  |  |  |  |
| Calgary | 4.87 | ref | 1.50 | ref | 83.06 | ref | 62.83 | ref |
| Edmonton | 1.08 | **0.22**<br>(0.19, 0.26) | 11.93 | **7.96**<br>(7.09, 8.94) | 81.06 | 0.99<br>(0.98, 1.00) | 51.92 | **0.83**<br>(0.82, 0.85) |
| Central | 24.73 | **5.10**<br>(4.72, 5.50) | 11.61 | **7.74**<br>(6.79, 8.83) | 78.31 | **0.96**<br>(0.94, 0.97) | 64.04 | 1.02<br>(1.00, 1.05) |
| North | 5.35 | 1.10<br>(0.97, 1.26) | 9.25 | **6.17**<br>(5.37, 7.08) | 79.42 | **0.97**<br>(0.96, 0.99) | 52.70 | **0.84**<br>(0.82, 0.87) |
| South | 3.98 | **0.82**<br>(0.69, 0.97) | 0.29 | **0.19**<br>(0.11, 0.35) | 78.65 | **0.96**<br>(0.94, 0.98) | 5.79 | **0.09**<br>(0.08, 0.11) |
| **School authority type** |  |  |  |  |  |  |  |  |
| Public | 6.56 | ref | 7.38 | ref | 80.85 | ref | 54.48 | ref |
| Catholic | 4.20 | **0.64**<br>(0.58, 0.70) | 6.10 | **0.83**<br>(0.76, 0.89) | 83.56 | **1.03**<br>(1.02, 1.04) | 57.24 | **1.05**<br>(1.03, 1.07) |
| Private | 1.73 | **0.26**<br>(0.19, 0.36) | 0.80 | **0.11**<br>(0.07, 0.16) | 54.78 | **0.68**<br>(0.65, 0.71) | 39.99 | **0.73**<br>(0.69, 0.78) |
| Charter | 0.31 | **0.05**<br>(0.02, 0.15) | 1.42 | **0.19**<br>(0.11, 0.32) | 90.02 | **1.11**<br>(1.09, 1.14) | 54.99 | 1.01<br>(0.95, 1.07) |
| Francophone | 3.61 | **0.55**<br>(0.37, 0.82) | 4.10 | **0.55**<br>(0.39, 0.79) | 82.37 | 1.02<br>(0.97, 1.07) | 53.93 | 0.99<br>(0.91, 1.08) |

*Notes.* CI = confidence interval; HPV = human papillomavirus; MenC-ACYW = meningococcal A, C, Y, W-135; ref = reference category; RR = relative risk.
Coverage values calculated as the percentage of children fully vaccinated (i.e., received at least two doses of HPV or one dose of MenC-ACYW by July 31 of each respective school year.
Bolded indicates significant difference from reference category.

#### Cumulative coverage

After removing those who died or left the province before the end of the follow-up period (September 1, 2021), 51,494 children from the 2019-2020 cohort were included in the cumulative coverage analysis. As shown in Figure 2, HPV full coverage in the 2019-2020 cohort improved over the approximately one-year follow-up period, from 5.6% on July 31, 2020 to 50.2% on September 1, 2021. However, in comparison to the 2017-2018 pre-pandemic cohort, HPV coverage remained significantly lower in the 2019-2020 pandemic cohort even after follow-up (50.2% vs. 66.4%; absolute difference: 16.2%; 95% CI: 15.6-16.8%; p < 0.001).

**Figure 2.**
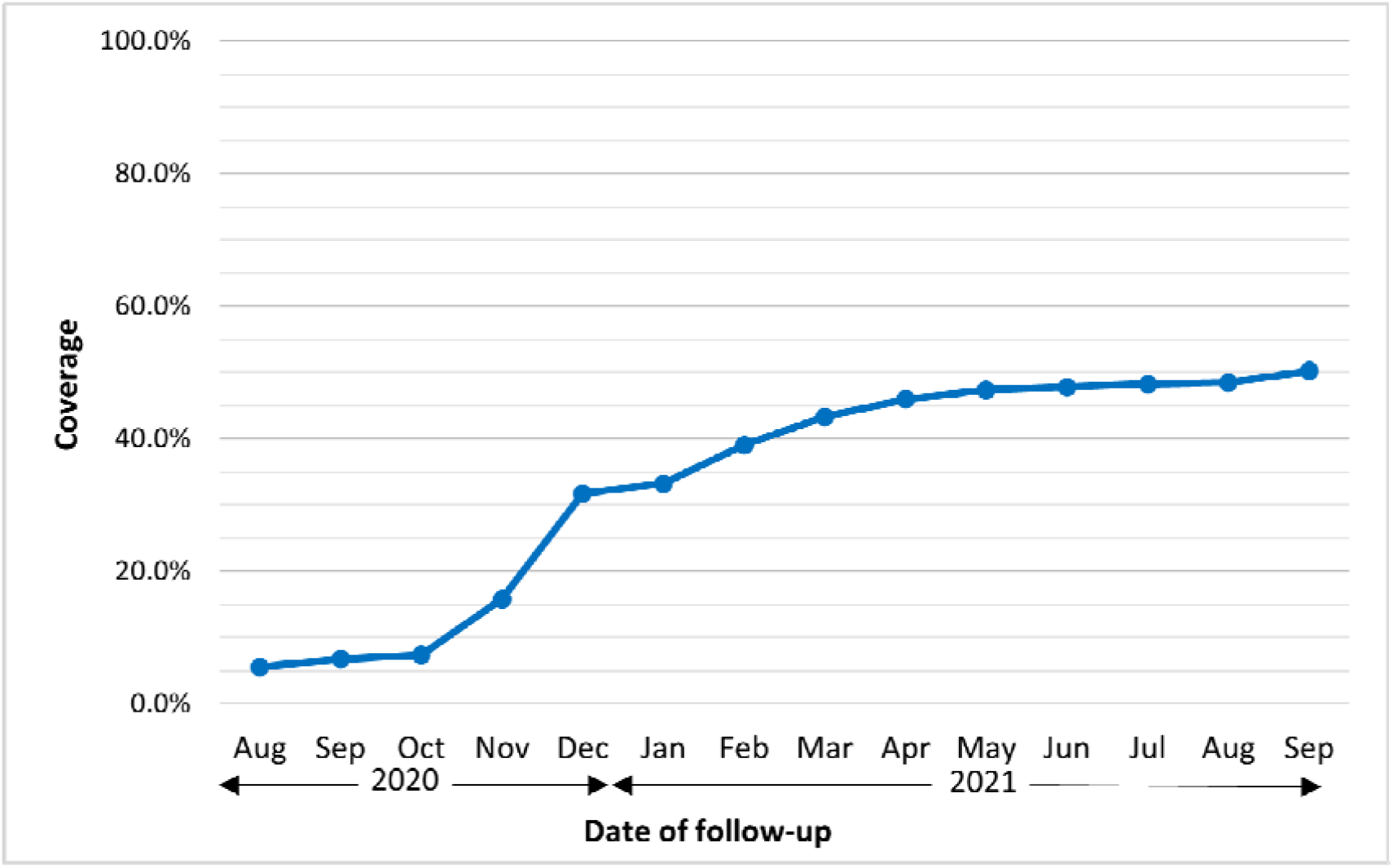
Cumulative coverage graph displaying HPV full coverage^a^ for grade 6 children from the 2019-2020 school cohort during the follow-up period (July 31, 2020 to September 1, 2021). *Notes*. HPV = human papillomavirus. ^a^Full coverage defined as the percentage of children who received all scheduled doses of HPV vaccine by the follow-up date (i.e., at least two doses).

When stratified by geographic health zone (Figure 3a), Central zone had the highest HPV coverage at the beginning of the follow-up period, but by December 2020 Calgary had surpassed Central zone and had the highest coverage until the end of follow-up. Most zones, other than South zone, had a sharp increase in coverage around October-December 2020, and steady increases in coverage throughout 2021. South zone did not display a large increase in coverage until the end of the follow-up period, but still had the lowest coverage.

**Figure 3.**
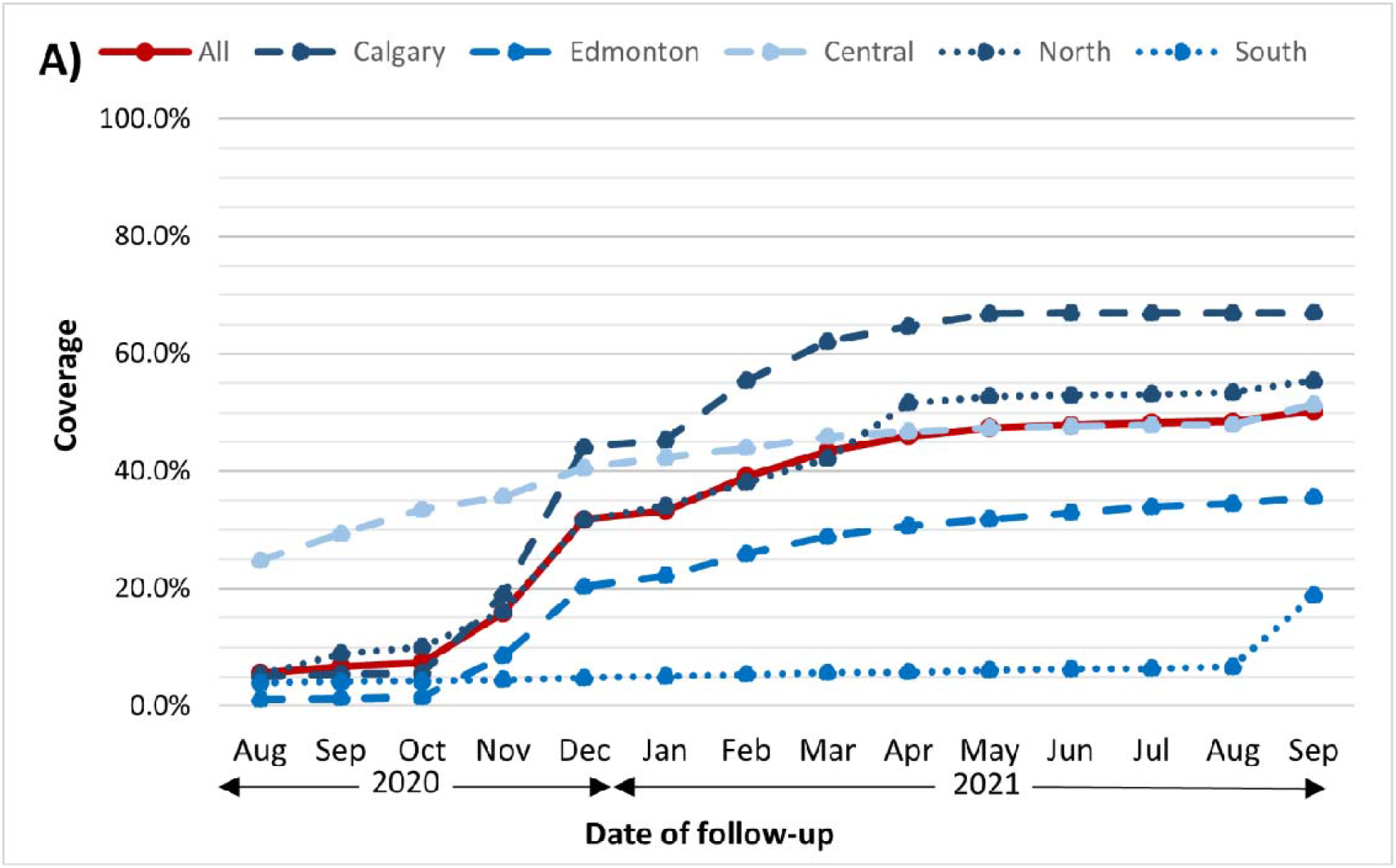
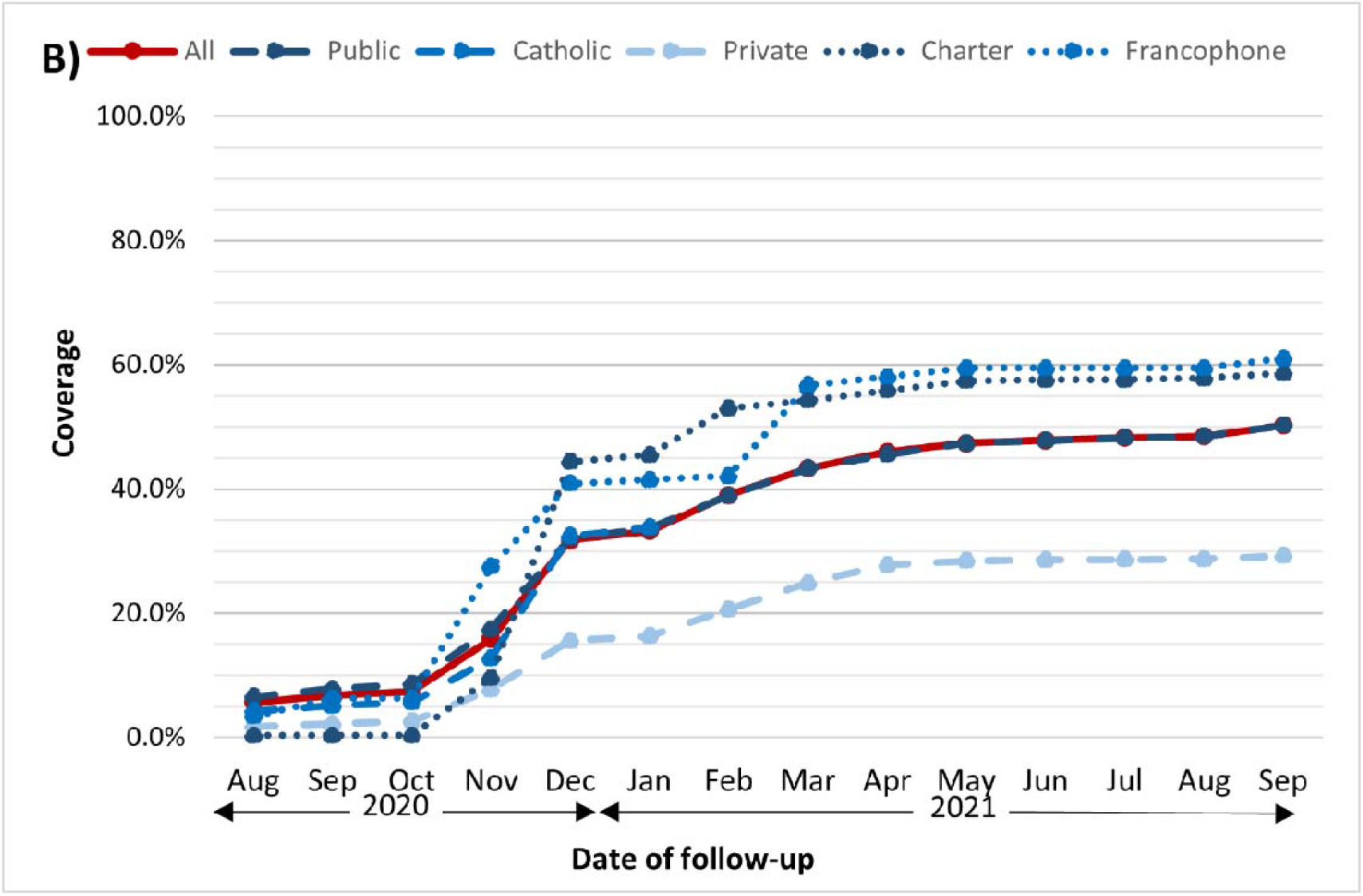
Cumulative coverage graph displaying HPV full coverage^a^ for grade 6 children from the 2019-2020 school cohort during the follow-up period (July 31, 2020 to September 1, 2021) stratified by school (a) geographic health zone (top) (b) authority type (bottom). *Notes*. HPV = human papillomavirus. ^a^Full coverage defined as the percentage of children who received all scheduled doses of HPV vaccine by the follow-up date (i.e., at least two doses).

Based on school authority type (Figure 3b), all had similar coverage for HPV throughout follow-up, except for private schools which had the lowest coverage from November 2020 to the end of follow-up. All school types exhibited a sharp increase in coverage around October-December 2020 and had steady increases in coverage throughout 2021. Francophone and charter schools had the highest HPV coverage at the end of follow-up.

### MenC-ACYW coverage

Full coverage (i.e., at least one dose) for the MenC-ACYW vaccine on July 31 of each respective school year was 86.8% in the 2017-2018 cohort, 80.7% in the 2019-2020 cohort, and 54.6% in the 2020-2021 cohort (Figure 4). In comparison to the 2017-2018 pre-pandemic cohort, coverage for MenC-ACYW was significantly lower in the 2019-2020 pandemic cohort (80.7% vs. 86.8%; absolute difference: 6.1%; 95% CI: 5.6-6.5%; p < 0·001) and the 2020-2021 pandemic cohort (54.6% vs. 86.8%; absolute difference: 32.2%; 95% CI: 31.6-32.7%; p < 0.001).

**Figure 4.**
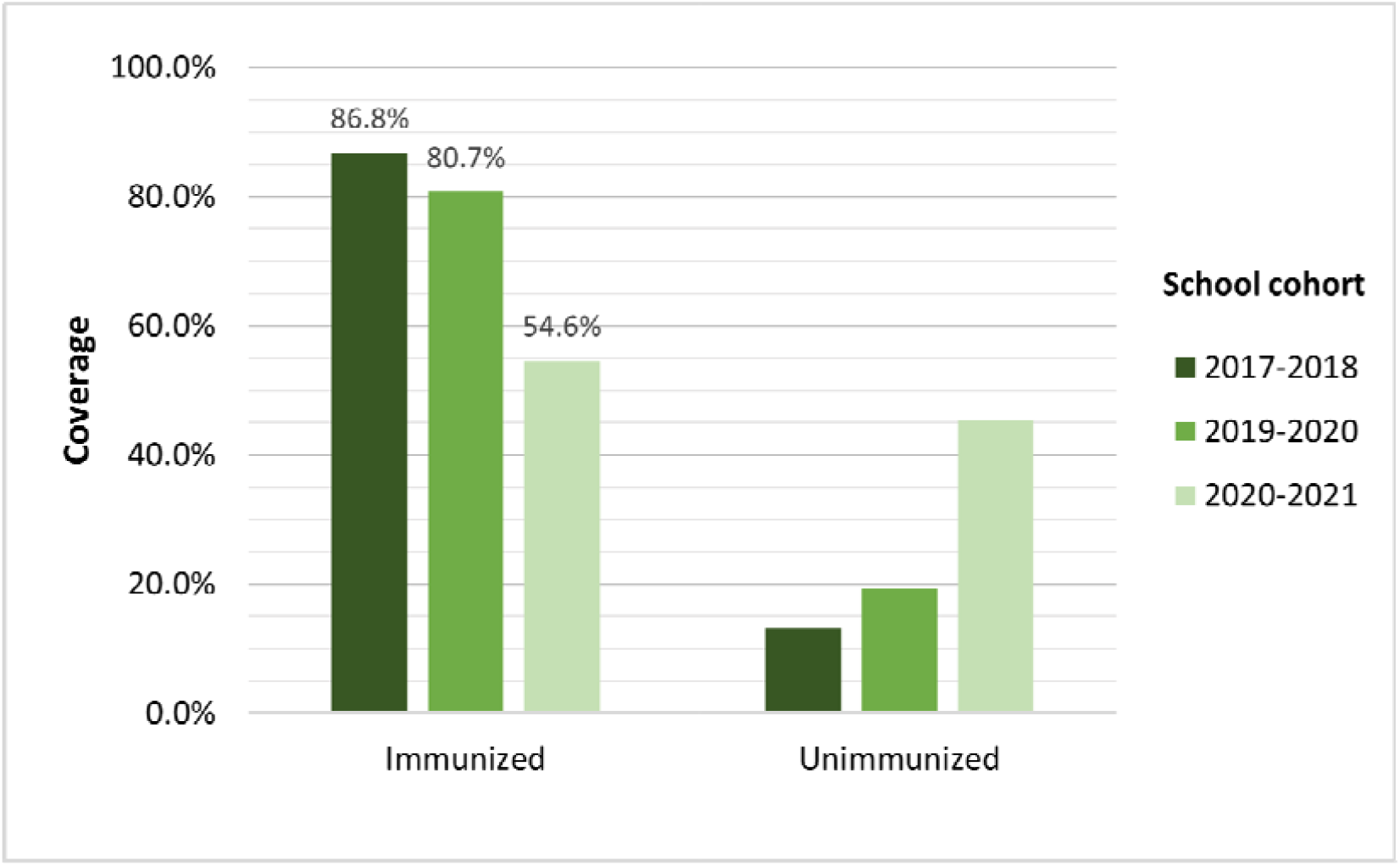
Coverage (immunized^a^, unimmunized) in the pre-pandemic (2017-2018) and pandemic (2019-2020, 2020-2021) school cohorts at July 31 of each respective school year for the MenC-ACYW vaccine. *Notes*. MenC-ACYW = meningococcal A, C, Y, W-135. The HPV school immunization program in Alberta changed from a three-dose schedule given in grade 5 to a two-dose schedule given in grade 6 in the 2018-2019 school year. As no immunization data were available during the 2018-2019 school year for HPV vaccine, 2017-2018 was also used as the pre-pandemic comparison year for MenC-ACYW vaccine to be consistent across both vaccines. ^a^Immunized defined as the percentage of children who received all scheduled doses of MenC-ACYW vaccine by July 31 of each respective school year (i.e., at least one dose).

MenC-ACYW coverage also differed by geographic health zone and school type, but slightly less than for HPV (Table 2). Across health zones, coverage ranged from 78.3% (Central) to 83.1% (Calgary) in 2019-2020 and 5.8% (South) to 64.0% (Central) in 2020-2021. When analyzed by school type, coverage was significantly lower in private schools in both the 2019-2020 and 2020-2021 school years (54.8% and 40.0%), and highest in charter schools in 2019-2020 (90.0%) and publicly-funded Catholic schools in 2020-2021 (57.2%).

#### Cumulative coverage

After removing those who died or left the province before the end of the follow-up period (September 1, 2021), 45,183 children from the 2019-2020 cohort were included in the cumulative coverage analysis. As shown in Figure 5, MenC-ACYW vaccine coverage in the 2019-2020 cohort slightly improved over the approximately one-year follow-up period, from 80.7% on July 31, 2020 to 83.0% on September 1, 2021. However, in comparison to the 2017-2018 pre-pandemic cohort, MenC-ACYW coverage remained significantly lower in the 2019-2020 pandemic cohort even after the one-year follow-up (83.0% vs. 86.8%; absolute difference: 3.7%; 95% CI: 3.3-4.2%; p < 0.001).

**Figure 5.**
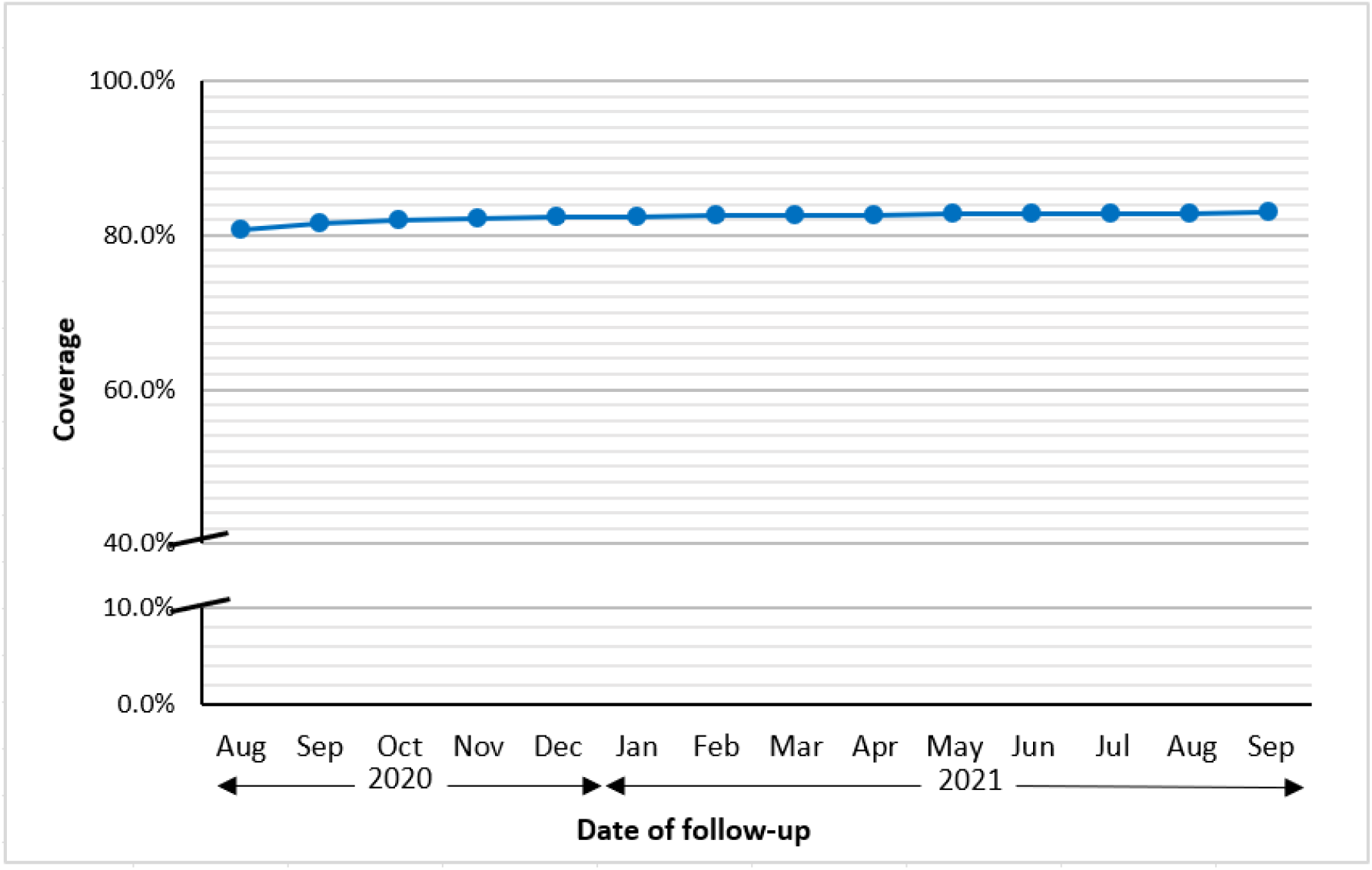
Cumulative coverage graph displaying MenC-ACYW coverage^a^ for grade 9 children from the 2019-2020 school cohort during the follow-up period (July 31, 2020 to September 1, 2021). *Notes*. MenC-ACYW = meningococcal A, C, Y, W-135. ^a^Coverage defined as the percentage of children who received all scheduled doses of MenC-ACYW vaccine by the follow-up date (i.e., at least one dose).

When stratified by geographic health zone (Figure 6a), all zones had steady MenC-ACYW coverage throughout the follow-up period, with Calgary having slightly higher coverage at the beginning of follow-up, and Edmonton having highest coverage at the end of follow-up.

**Figure 6.**
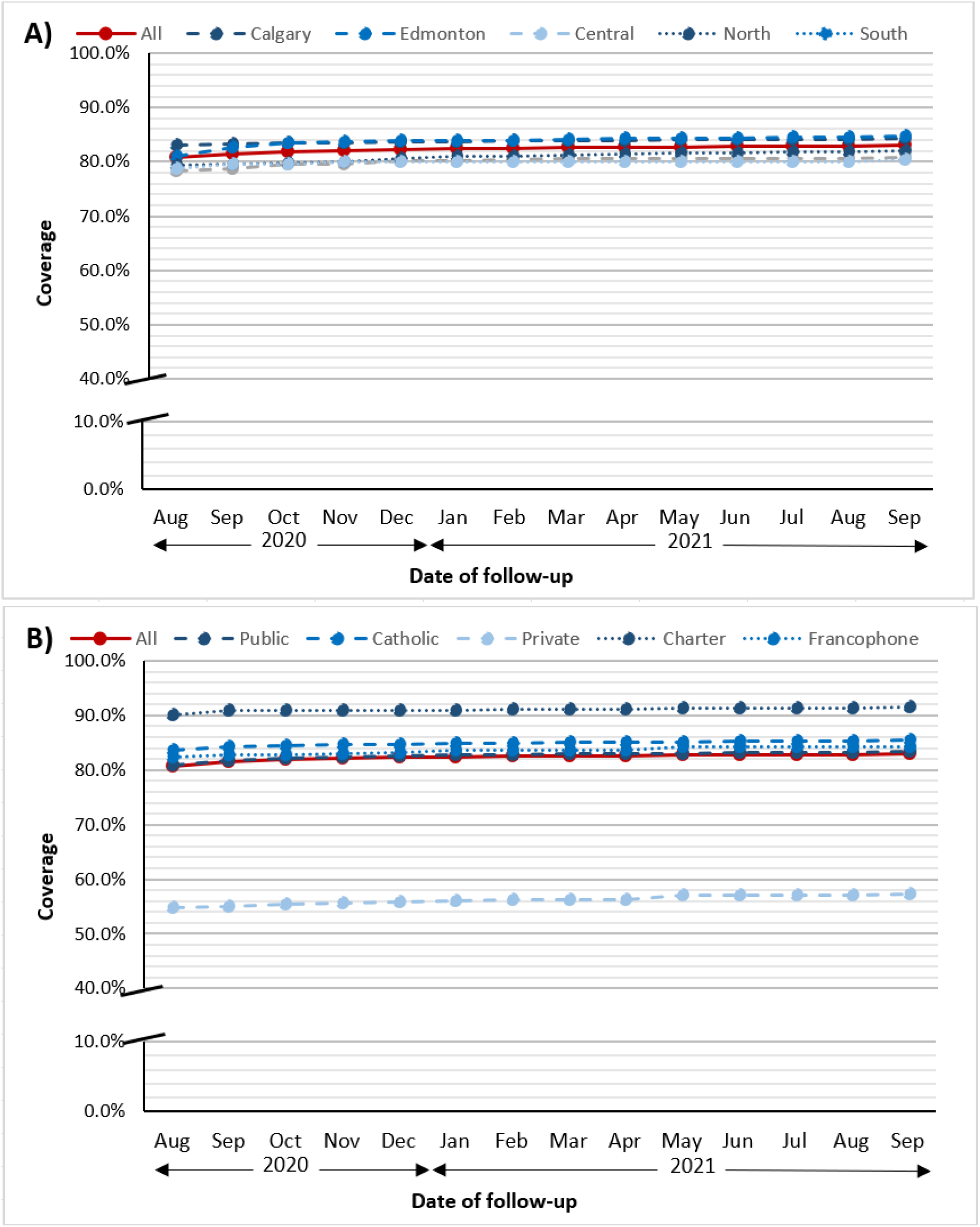
Cumulative coverage graph displaying MenC-ACYW coverage^a^ for grade 9 children from the 2019-2020 school cohort during the follow-up period (July 31, 2020 to September 1, 2021) stratified by school a) geographic health zone (top) b) authority type (bottom). *Notes*. MenC-ACYW = meningococcal A, C, Y, W-135. ^a^Coverage defined as the percentage of children who received all scheduled doses of MenC-ACYW vaccine by the follow-up date (i.e., at least one dose).

Based on school authority type (Figure 6b), private schools had the lowest coverage for MenC-ACYW throughout the entire follow-up period, while charter schools had the highest. Public, publicly-funded Catholic, and francophone schools had similar coverage during follow-up. Coverage was steady for all school types throughout the follow-up period.

## Discussion

The COVID-19 pandemic has significantly impacted health care systems worldwide, including routine immunization programs. We found that HPV and MenC-ACYW immunization coverage was significantly lower in cohorts of schoolchildren who were eligible for immunization during the pandemic in comparison to before the pandemic. This decline in coverage seen during the pandemic is consistent with previous literature on other routine immunizations internationally, including infant,^3,15,16^ preschool,^3,15^ and adolescent.^2,3,17,18^

Coverage for HPV was below that of MenC-ACYW. This finding is likely explained by the fact that MenC-ACYW is a single-dose vaccine, so it was easier to complete this series before the COVID-19-related school closures, whereas the HPV series requires two doses, six months apart. Many students in both pandemic cohorts had received a single dose of HPV by the end of the school year (71.5% in the 2019-2020 and 54.5% in the 2020-2021 cohort), therefore additional follow-up efforts are needed to ensure that these partially immunized children complete the series to maximize protection against HPV.

Coverage also differed by geographic health zone. Discrepancies in coverage across health zones likely reflects the epidemiology of COVID-19 in each region and the available resources to provide immunizations to schoolchildren. For example, in South zone school immunizations were delayed until the end of July 2021 due to the COVID-19 pandemic response, which may explain the lower coverage in this zone.^19^ On the other hand, personal communication with public health officials in Central zone, which had higher HPV coverage (24.7%) than all other zones at the end of the 2019-2020 school year, revealed that additional immunization clinics were held at community health centres throughout June-August 2020 to catch-up students with missed doses, particularly in smaller rural sites that were not overwhelmed by COVID-19 testing.

Similarly, private schools had low coverage for all vaccines in both school years in comparison to the other school types. Previous literature from Canada and the U.S. has found that private schools have lower immunization coverage even prior to the pandemic.^20,21^ The differing coverage across geographic health zones and types has important implications for catch-up planning and implementation. Stratifying coverage by health zone and type may be useful for identifying where to focus catch-up immunization programs.

The cumulative coverage analysis found that coverage for both HPV and MenC-ACYW improved over a one-year period in individuals from the 2019-2020 cohort who missed immunizations in the first year of the pandemic, although the improvement was not as large (or necessary) for MenC-ACYW. For HPV, sharp increases in coverage were seen throughout October-December 2020, suggesting the implementation of fall catch-up programs. Although this is an encouraging finding, coverage still remained below pre-pandemic levels at the end of follow-up, necessitating further catch-up for this group. The 2020-2021 cohort had low coverage for both HPV and MenC-ACYW vaccines on July 31, 2021, meaning that catch-up for this group is urgently needed.

### Strengths and limitations

To our knowledge, this study is one of the first to assess how adolescent immunization coverage has changed over time during the COVID-19 pandemic and whether it has returned to pre-pandemic levels. A strength of our study was the use of a population-based repository that contains complete and timely information on all childhood immunizations, as well as complete denominator data. Data were included over a one-year period to determine whether coverage was recovering to pre-pandemic levels, which was previously unknown.

A limitation is that there was a change in programming for the HPV vaccine during the 2018-2019 school year, moving from three doses in grade 5 to two doses in grade 6. As there were no HPV immunizations provided in this school year, we could not use this year for comparison. This meant that our definition of fully vaccinated was three or more doses for the 2017-2018 cohort, but two or more doses for the 2019-2020 and 2020-2021 cohorts. However, the percentage of individuals with two or more doses of HPV vaccine in the 2017-2018 cohort was 74.3%, so our difference in coverage estimates based on three dose coverage are conservative and with the same direction of effect (i.e., coverage is significantly lower in the pandemic cohorts in comparison to the pre-pandemic cohort). Additionally, as this study focused on immunization coverage for Albertan school children, findings may not be generalizable to other jurisdictions. Finally, as this is a descriptive analysis, we cannot be certain whether the differences in coverage seen across geographic health zones and school type can be attributed to individual, school, or zone-level factors. Further analyses are needed to understand the relationship between these factors and immunization coverage during the COVID-19 pandemic.

## Conclusion

Routine childhood immunizations are important to prevent outbreaks of vaccine-preventable diseases, which have occurred within the past decade. Additionally, individuals who miss immunizations at the scheduled time may be less likely to catch-up later.^5^ In this study, coverage analyses showed that coverage for HPV and MenC-ACYW vaccines in cohorts of school children eligible for immunization during the COVID-19 pandemic declined in comparison to the pre-pandemic period. Although our findings show that some catch-up has occurred, additional efforts will be needed to ensure children are fully protected. This will require a multi-pronged approach, utilizing existing strategies such as holding additional catch-up clinics at public health centres or schools, as well as possible new strategies including providing missing routine vaccine doses at other vaccine appointments (e.g., COVID-19, influenza)^22,23^ or any contact with the health system (e.g., visits at primary care practices, pharmacies, or hospitals).^24^ To date, the 2020-2021 cohort had the lowest immunization coverage, suggesting that this group may be a priority for catch-up programs.

## Data Availability

The steward of the data used in this study is the Alberta Ministry of Health, who maintains the data for the purpose of health system administration. Thus, the authors are not at liberty to make the data publicly available.

## Abbreviations

AHCIP: Alberta Health Care Insurance Plan
CI: confidence interval
COVID-19: coronavirus disease 2019
HPV: human papillomavirus
Imm/ARI: Immunization and Adverse Reaction to Immunization
MenC-ACYW: meningococcal conjugate A, C, Y, W-135
PSIR: Provincial School Immunization Record
SD: standard deviation.

## Acknowledgments

This work was part of a larger project conducted by the COVImm study team.

## Contributors

Hannah Sell: Conceptualization, methodology, data curation, software, formal analysis, investigation, visualization, writing – original draft and review & editing; Yuba Raj Paudel: Conceptualization, methodology, software, data curation, writing - review and editing, supervision; Donald Voaklander: Conceptualization, methodology, supervision, writing – review and editing; Shannon MacDonald: Conceptualization, methodology, funding acquisition, supervision, writing – review and editing.

## References

1. WHO and UNICEF warn of a decline in vaccinations duing COVID-19. World Health Organization. Published July 15, 2020. Accessed June 22, 2021. https://www.who.int/news/item/15-07-2020-who-and-unicef-warn-of-a-decline-in-vaccinations-during-covid-19

2. O’Leary ST, Trefren L, Roth H, Moss A, Severson R, Kempe A. Number of childhood and adolescent vaccinations administered before and after the COVID-19 outbreak in Colorado. JAMA Pediatr. 2021;175(3):305–7. doi: 10.1001/jamapediatrics.2020.4733.

3. DeSilva MB, Haapala J, Vazquez-Benitez G, et al. Association of the COVID-19 pandemic with routine childhood vaccination rates and proportion up to date with vaccinations across 8 US health systems in the Vaccine Safety Datalink. JAMA Pediatr. 2022;176(1):68–77. doi: 10.1001/jamapediatrics.2021.4251.

4. COVID-19 orders and legislation. Alberta Government. Updated February 28, 2022. Accessed April 26, 2022. https://www.alberta.ca/covid-19-orders-and-legislation.aspx

5. National Advisory Committee on Immunization. Interim guidance on continuity of immunization programs during the COVID-19 pandemic. Government of Canada. Updated May 13, 2020. Accessed April 1, 2022. https://www.canada.ca/en/public-health/services/immunization/national-advisory-committee-on-immunization-naci/interim-guidance-immunization-programs-during-covid-19-pandemic.html

6. Guidance for immunization services during COVID-19. Ontario Ministry of Health. Published August 26, 2020. Updated October 28, 2021. Accessed April 26, 2022. Available from: https://www.health.gov.on.ca/en/pro/programs/publichealth/coronavirus/docs/Immunization_Services_during_COVID-19_08-26-2020.pdf

7. Sell H, Assi A, Driedger SM, et al. Continuity of routine immunization programs in Canada during the COVID-19 pandemic. Vaccine. 2021;39(39):5532–7. doi: 10.1016/j.vaccine.2021.08.044.

8. Stick to immunization schedule during the COVID-19 pandemic, paediatricians urge. Canadian Pediatric Society. Published April 30, 2020. Accessed April 1, 2022. https://www.cps.ca/en/media/stick-to-immunization-schedule-during-the-covid-19-pandemic

9. Economic dashboard. Alberta Government. Updated March 2022. Accessed April 26, 2022. https://economicdashboard.alberta.ca/

10. AHS map and zone overview. Alberta Health Services. Published 2021. Accessed April 26, 2022. https://www.albertahealthservices.ca/assets/about/publications/ahs-ar-2021/zones.html

11. Immunization - school services. Alberta Health Services. Accessed April 26, 2022. https://www.albertahealthservices.ca/findhealth/service.aspx?id=4209

12. Interactive Health Data Application. Alberta Government. Accessed April 26, 2022. http://www.ahw.gov.ab.ca/IHDA_Retrieval/ihdaData.do

13. MacDonald SE, Russell ML, Liu XC, et al. Are we speaking the same language? an argument for the consistent use of terminology and definitions for childhood vaccination indicators. Hum Vaccin Immunother. 2019;15(3):740–7. doi: 10.1080/21645515.2018.1546526.

14. Human Papillomavirus 9-Valent vaccine biological page. Alberta Health Services. Updated January 1, 2021. Accessed April 26, 2022. https://www.albertahealthservices.ca/assets/info/hp/cdc/If-hp-cdc-hpv-bio-pg-07-241.pdf

15. Bramer CA, Kimmins LM, Swanson R, et al. Decline in child vaccination coverage during the COVID-19 pandemic — Michigan Care Improvement Registry, May 2016-May 2020. Morb Mortal Wkly Rep. 2020;69(20):630–1. doi: 10.15585/mmwr.mm6920e1.

16. McDonald HI, Tessier E, White JM, et al. Early impact of the coronavirus disease (COVID-19) pandemic and physical distancing measures on routine childhood vaccinations in England, January to April 2020. Euro Surveill. 2020;25(19):1–6. doi: 10.2807/1560-7917.ES.2020.25.19.2000848.

17. Murthy BP, Zell E, Kirtland K, et al. Impact of the COVID-19 pandemic on administration of selected routine childhood and adolescent vaccinations — 10 U.S. jurisdictions, March-September 2020. Morb Mortal Wkly Rep. 2021;70(23):840–5. doi: 10.15585/mmwr.mm7023a2.

18. Saxena K, Marden JR, Carias C, et al. Impact of the COVID-19 pandemic on adolescent vaccinations: projected time to reverse deficits in routine adolescent vaccination in the United States. Curr Med Res Opin. 2021;37(12):2077–87. doi: 10.1080/03007995.2021.1981842.

19. Routine school immunizations to resume in July in South zone. Alberta Health Services. Published July 22, 2021. Accessed February 9, 2022. https://www.albertahealthservices.ca/news/releases/2021/Page16065.aspx

20. Carpiano RM, Bettinger JA. Vaccine coverage for kindergarteners: Factors associated with school and area variation in Vancouver, British Columbia. Vaccine Rep. 2016;6:50–5. doi:10.1016/j.vacrep.2016.10.001.

21. Lai YK, Nadeau J, McNutt LA, Shaw J. Variation in exemptions to school immunization requirements among New York State private and public schools. Vaccine. 2014;32(52):7070–6. doi:10.1016/j.vaccine.2014.10.077.

22. Interim clinical considerations for use of COVID-19 vaccines currently approved or authorized in the United States. Centers for Disease Control and Prevention. Updated April 21, 2022. Accessed April 26, 2022. https://www.cdc.gov/vaccines/covid-19/clinical-considerations/covid-19-vaccines-us.html?CDC_AA_refVal=https%3A%2F%2Fwww.cdc.gov%2Fvaccines%2Fcovid-19%2Finfo-by-product%2Fclinical-considerations.html#Coadministration

23. National Advisory Committee on Immunization. Recommendations on the use of COVID-19 vaccines. Government of Canada. Published October 22, 2021. Accessed January 20, 2022. https://www.canada.ca/en/public-health/services/immunization/national-advisory-committee-on-immunization-naci/recommendations-use-covid-19-vaccines.html

24. MacDonald NE, Comeau J, Dubé E, Bucci L, Graham JE. A public health timeline to prepare for COVID-19 vaccines in Canada. Can J Public Health. 2020;111(6):945–52. doi: 10.17269/s41997-020-00423-1.

