## Supplemental tables and figures for "School immunization coverage during the COVID-19 pandemic: A retrospective cohort study"

**Appendix**

**Table A1**. Number and timing of doses for school-based vaccines in Alberta, Canada.

| **Vaccine** | **Number and timing of doses** | **Minimum interval between doses** |
| --- | --- | --- |
| Hepatitis B | 2019-2020 school year and beyond:  2 doses in Grade 6  2017-2018 school year and prior:  3 doses in Grade 5 | Minimum interval between doses 1 & 2: 6 months  2^nd^ dose: 1 month after dose 1  3^rd^ dose: 6 months after dose 1 |
| Human papillomavirus (HPV) | 2018-2019 school year and beyond:  2 doses in Grade 6  (3 doses if first dose is at age 15 years or older)  2017-2018 school year and prior:  3 doses in Grade 5 | Minimum age dose 1: 9 years  Minimum interval between doses 1 & 2: 6 months    Minimum age dose 1: 9 years  2^nd^ dose: 2 months after dose 1  3^rd^ dose: 4 months after dose 1 |
| Diphtheria-tetanus-acellular pertussis (dTap) | 1 dose in Grade 9 | Minimum age: 12 years  Minimum interval: N/A |
| Meningococcal Conjugate A, C, Y, W-135 (MenC-ACYW) | 1 dose in Grade 9 | Minimum age: 12 years  Minimum interval: N/A |

*Notes.* dTap: diphtheria-tetanus-acellular pertussis; HPV: human papillomavirus; MenC-ACYW: meningococcal conjugate A, C, Y, W-135; N/A: Not Applicable.

Sources: (Alberta Government, 2021e; Alberta Health Services, 2021b, 2021c).

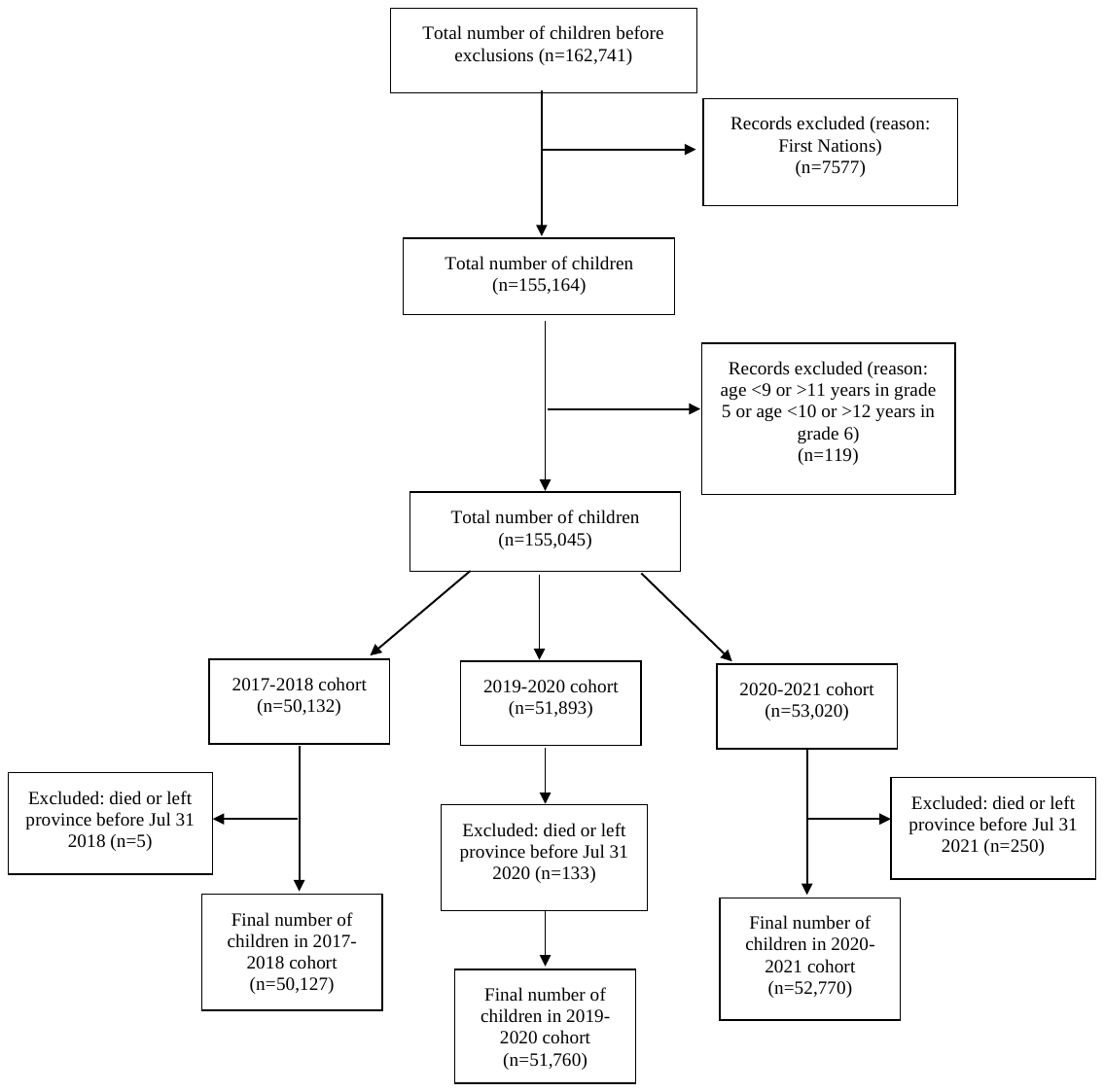

**Figure A1.** Flowchart of exclusion criteria for the HPV immunization cohorts for the 2017-2018, 2019-2020, and 2020-2021 school years.

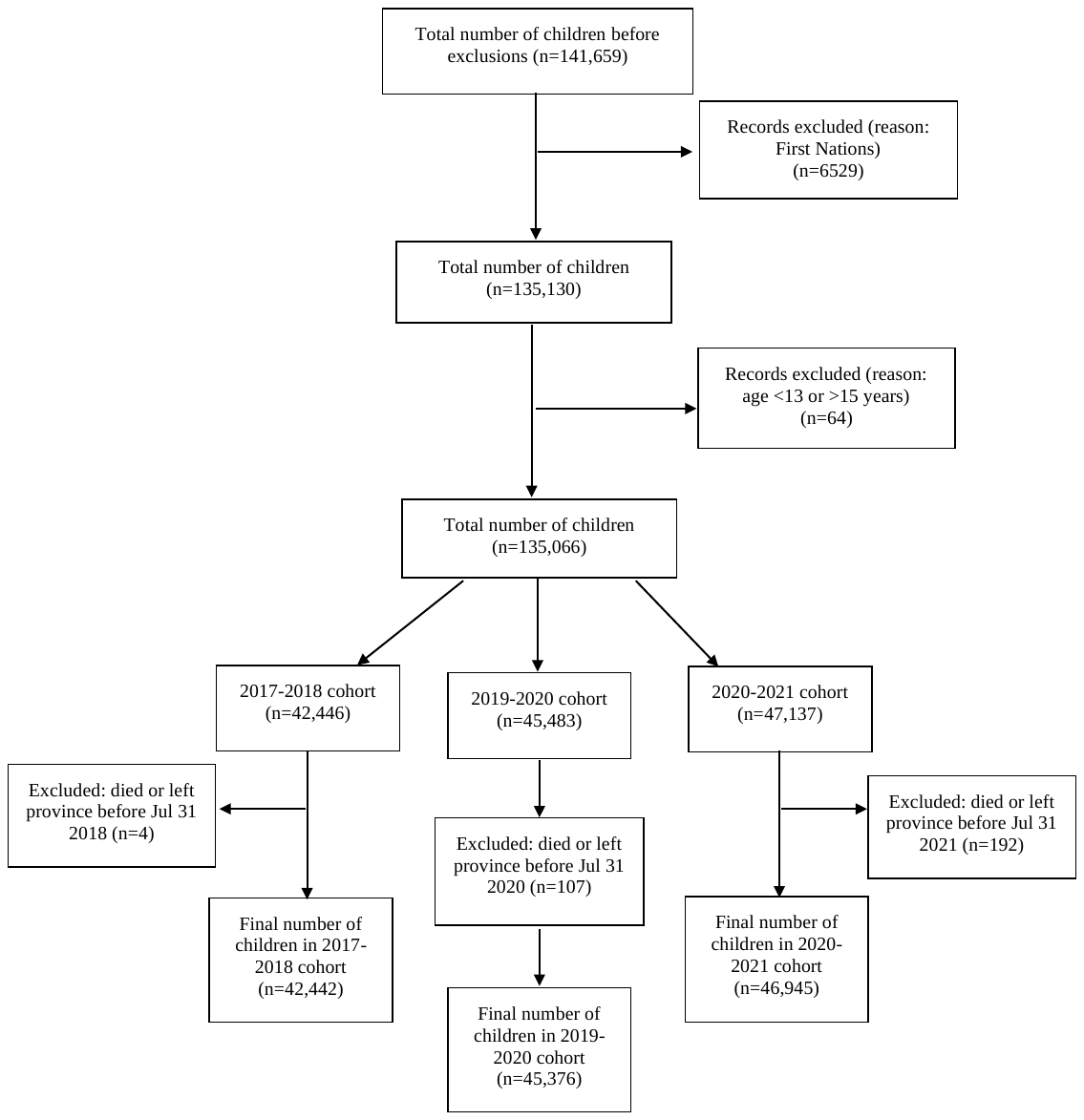

**Figure A2.** Flowchart of exclusion criteria for MenC-ACYW immunization cohorts for the 2017-2018, 2019-2020, and 2020-2021 school years.
